# The Case Against the ‘S’: Is Functional Neurological Disorder(s) One Condition or Many?

**DOI:** 10.64898/2026.03.19.26348846

**Authors:** David D.G. Palmer, Mark J. Edwards, Jason B. Mattingley

## Abstract

**Background:** Functional neurological disorder (FND) is one of the most common, but least researched, conditions in neurology. Debate exists as to whether the clinical entity referred to as FND is truly a single disorder or is in fact multiple entities which have been erroneously amalgamated into the same condition. We sought to provide empirical evidence on this question by treating it as a problem of model comparison.

**Methods:** We formulated statistical models equivalent to: (1) FND being a single entity with variation in phenotype, represented by latent trait (binary factor/item response theory) models, and (2) FND being multiple discrete entities, represented by latent class analysis (LCA) models. We fitted these models to data on the symptoms experienced by 697 people with FND from the FND Research Connect database (fnd-research.org) and used Bayesian model comparison methods to compare them.

**Results:** All but one of the latent trait models, representing FND as a single entity with heterogeneous phenotype, fit the data better than all the LCA models. Secondary analysis of the LCA models showed results compatible with the models capturing discretisation of continuous variation rather than true discrete categories.

**Discussion:** Our results suggest that the symptom structure of FND is the result of a single pathophysiological process, either as a single entity, or a common pathway preceded by multiple causative processes where the common pathway is solely responsible for the phenotype of the condition.

## Introduction

Functional neurological disorder (FND) is among the most common and intriguing conditions encountered in neurological practice.^1^ Despite its prevalence, and increasing research interest in the condition, fundamental questions about its underlying nature remain unanswered. Perhaps the most basic of these is whether FND is best understood as a single disorder with heterogeneous manifestations, or as a collection of distinct but clinically overlapping conditions that are currently grouped under a common diagnostic label.

The question of whether there is one or many FND(s) is reflected not only in the academic literature, but also in clinical discourse, where it is variably referred to in the singular or plural. Such linguistic variation is not merely semantic. Competing views of FND as one condition or many imply different assumptions about aetiology, prognosis, and approach to treatment, and they shape the way scientific studies are designed and interpreted. If FND comprises multiple distinct conditions, then aggregating patients into a single diagnostic category risks obscuring meaningful differences and diluting true effects. Conversely, if FND represents a single pathophysiological process with variable expression, then any adequate account of its pathophysiology must also explain its apparent heterogeneity.

Much of the existing debate has been informed by conceptual models, which emphasise either shared neurobiological processes across functional presentations or heterogeneity between symptom-defined subtypes.^2,3^ While such approaches are essential, mechanistic arguments alone do not resolve questions of diagnostic structure. Distinct pathophysiological processes may converge on similar clinical phenotypes, while a single underlying mechanism may give rise to diverse manifestations. In either case, differences in mechanism would be expected to leave some trace at the level of observable phenotype, even if these differences are subtle and only detectable at the population level.

We identify three broad possibilities for the structure of FND as an entity. First, FND might represent a single pathophysiological process which can express itself in various combinations of symptoms. An example of this situation could be considered that of a cerebral abscess: the pathology is the same wherever the abscess forms, but sensitivity to initial conditions (e.g. where the initial inoculum is deposited) mean that it might cause different symptoms depending on its location. A second possibility is that FND represents multiple pathologies which affect a single common pathway, thereby causing an identifiable syndrome despite the multitude of possible causes, but with heterogeneity which is attributable to the differences in underlying pathophysiologies. Parkinson’s disease is currently understood in this way, as a condition resulting from multiple pathologies, some of which are known (e.g. certain genetic mutations).^4^ The involvement of a common pathway by these pathologies means that the disease has an identifiable phenotype, but differences in underlying pathology may also be apparent based on the phenotype of the condition. A third possibility involves multiple processes which cause related phenotypes, but which share no mechanism. Optic neuritis might be considered an example of this: in recent decades, myelin oligodendrocyte glycoprotein (MOG) antibody-associated disease and aquaporin-4 associated neuromyelitis optica have been distinguished as separate entities from idiopathic and multiple sclerosis-associated optic neuritis.

From this perspective, the question of whether FND is one disorder or many can be framed as a problem of model comparison. The first scenario above could be represented by a class of models that posits a single latent disease process which produces a heterogeneous but structured distribution of symptoms across individuals, while the second and third could be represented by models of multiple latent disease processes which give rise to overlapping symptom profiles that are difficult to distinguish clinically. Epidemiological analysis of symptom patterns provides a way to evaluate the relative plausibility of these competing models without presupposing the nature or number of underlying mechanisms.

Symptom-based approaches have a long history in neurology and psychiatry as tools for clarifying disease structure, particularly where biomarkers are absent or incomplete. Demonstrating that symptom distributions are consistent with a single population, or alternatively that they show evidence of multiple separable subpopulations, places meaningful constraints on theoretical models and can inform subsequent mechanistic and interventional research.

Here we use epidemiological analysis of symptom data from a large cohort of individuals with FND to examine whether the observed pattern of symptoms is more consistent with a single heterogeneous disorder or with multiple latent disease processes. By comparing models that differ in their assumptions about underlying disease structure, we aim to contribute empirical evidence to a debate that has thus far been driven largely by theory and clinical intuition.

## Methods

In contrasting the two candidate nosological structures for FND—either that it represents a single entity, or that it is constituted by more than one underlying condition—we considered each to be represented by a distinct class of model. The possibility that the condition is a single entity with dimensionally varying phenotype, is represented by a latent trait model with traits, representing the number of dimensions along which phenotype varies. We contrasted this with a latent class model, which assumes the existence of discrete categories (disease entities), each of which generates its own pattern of symptoms. The case of multiple causal processes which share a single-common pathway could be represented by either of these models, depending on the relative contributions to symptom generation of the shared and unique parts of the pathophysiology (FFigure 1).

**Figure 1:**
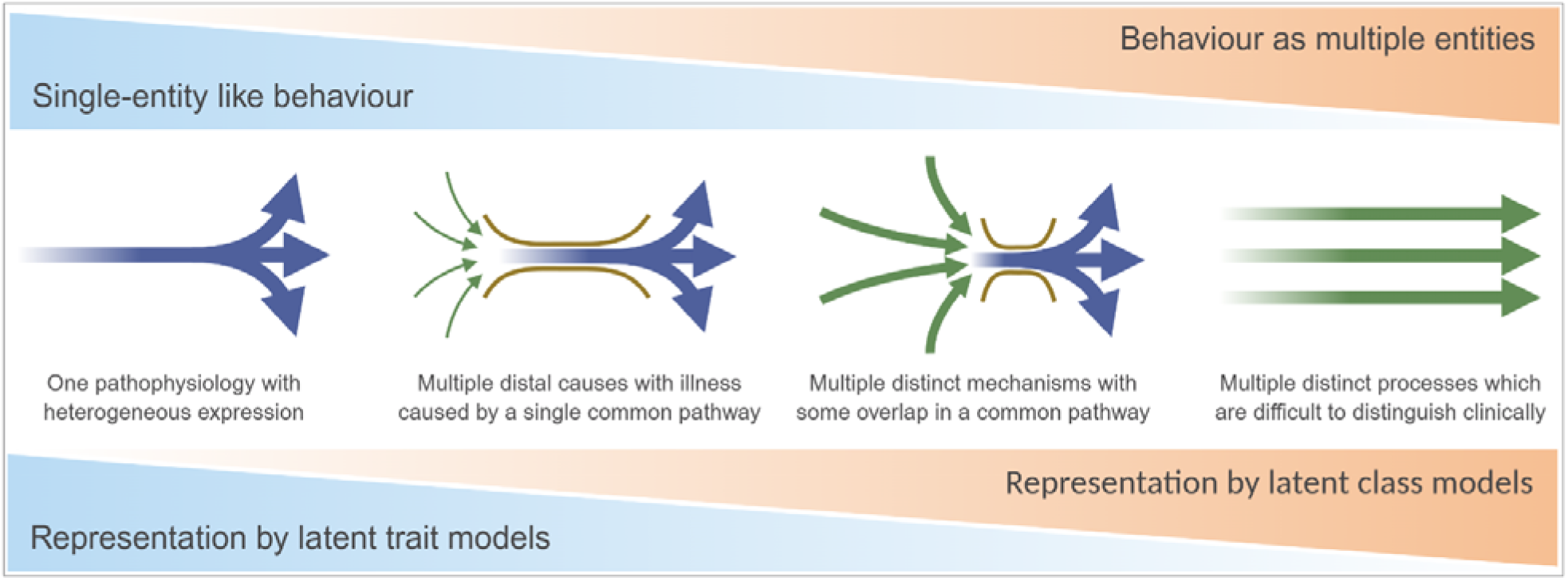
Schematic representation of the continuum of possible causative processes for FND and their correspondence to our statistical models.

We used data from FND Research Connect (fnd-research.org), an online research-recruitment platform and longitudinal data collection project for people with FND and controls. Data for which of 19 symptoms of FND had been experienced in the preceding two months by 697 participants with FND were used for this study. Data analysis was performed in Python, using the Stepmix package for specification and fitting of LCA models,^5^ and PyMC for specification and fitting of latent trait models.^6^ We created LCA models for class numbers from 1 to 8. Since latent trait models are more computationally costly to fit, we sequentially evaluated models with increasing numbers of latent traits until the addition of a further trait worsened model fit as assessed by K-fold cross-validated (CV) mean log predictive density, therefore ending at a 5-trait model.

Models were compared using patient-level K-fold cross-validated mean log predictive density. To examine the possibility that the identified classes of our LCA models approximated discretisations of a continuous latent structure rather than discrete subgroups, we additionally examined the following parameters: (1) effective number of classes, K_eff_, defined as the patient-level mean perplexity; (2) posterior membership uncertainty (boundary mass), defined as the proportion of participants for whom the posterior probability of their being a member of their most probable class was less than 0.8; and (3) low-dimensional embeddings of symptom similarity for these models as an aid to geometrical interpretation. These analyses were performed to aid interpretation of the latent class solutions and were not used for model selection.

To infer the qualitative appearances of the dimensions characterised by the best-fitting latent trait model, we post-processed the factors to maximise interpretability by performing a varimax rotation, then examined and descriptively summarised each by the symptoms with the greatest change in probability associated with movement along the dimension.

Data and analysis scripts are available through the Online Science Framework at (https://doi.org/10.17605/OSF.IO/CAXGF).

## Results

The data of 684 participants in the FND Research Connect database were used. The median age of included participants was 44 years (IQR 33 – 53). 86% were of female sex. 82% identified their gender as female, 14% as male, 3% as transgender, and 2% as non-binary.

Model fits as described by mean patient-level cross-validated log predictive density are presented in Figure 2. The difference in log predictive density between the best fitting (continuous) latent trait model (orange function) and the best fitting (discrete) latent class model (blue function) was 0.12, corresponding to an approximately 13% increase in predictive probability. All but the 1-dimensional latent trait models fitted the data better than the best fitting latent class model.

**Figure 2:**
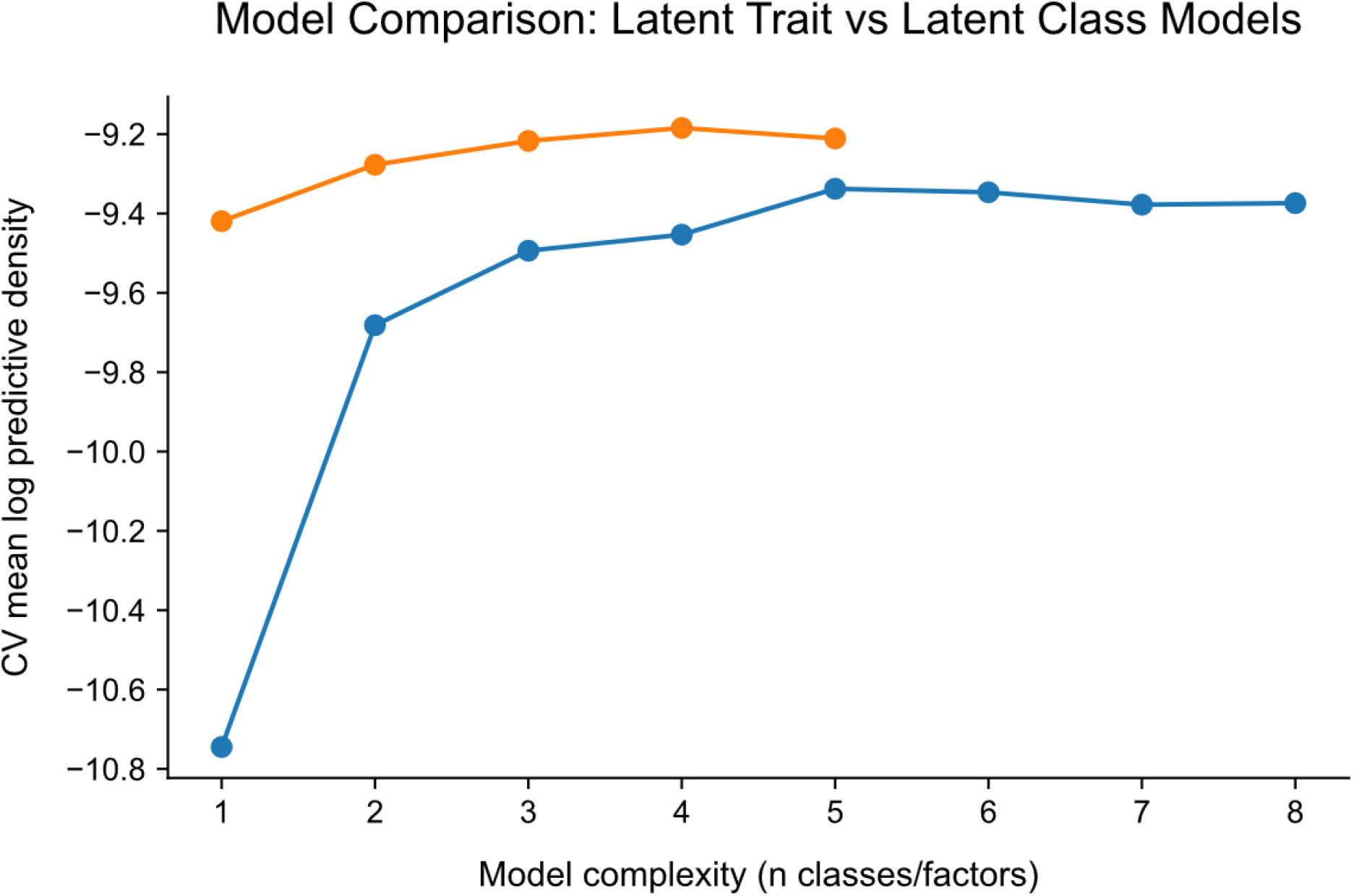
Mean patient-level cross-validated log predictive density for continuous latent trait models (orange) and discrete latent class models (blue). Continuous latent trait models provide superior out-of sample predictive performance, indicating that symptom distributions are better described by graded latent dimensions than discrete subtypes. The difference of 0.12 between the best fitting continuous model and the best fitting discrete model is equal to an approximately 13% increase in predictive probability.

To assess whether the classes identified by our LCA models represented true discrete classes or discretisations of underlying continuous variation, we performed further analysis on the best-fitting LCA model, which had 5 classes. For this model, the effective number of classes, K_Eff_, was 1.38, indicating that each patient’s data occupied a mean of 1.38 classes. Boundary mass was substantial, with 22.4% of patients having maximum posterior probability for class membership less than 0.8. To illustrate the latent class structure implied by this model, two dimensional embeddings of the Jaccard distance between 19-dimensional patient symptom profiles are presented in Figure 3. The embedding demonstrates substantial overlap between categories, with no appreciable gaps, compatible with the interpretation that the categories represent graded variation along continuous latent dimensions.

**Figure 3:**
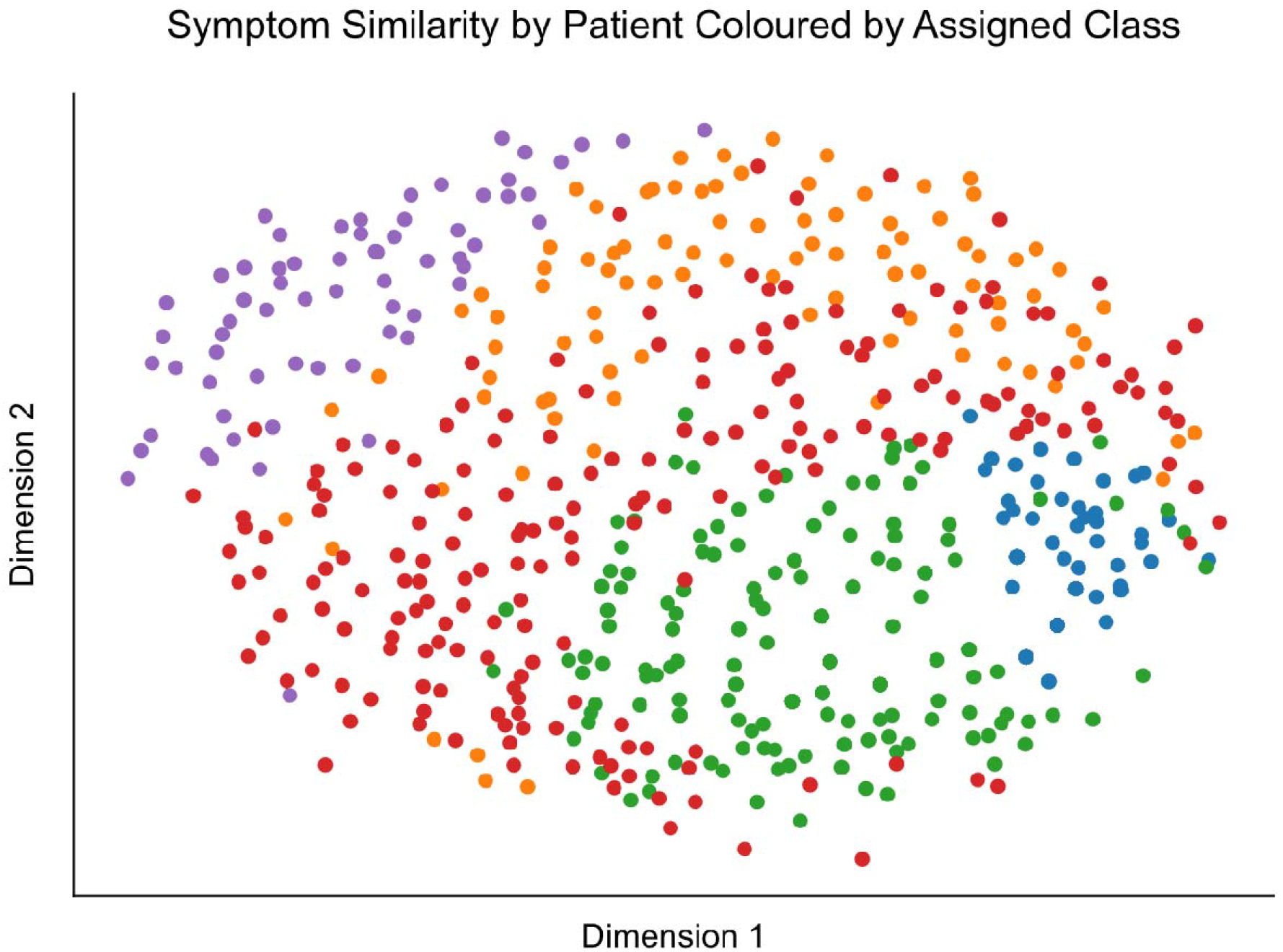
Two-dimensional embedding of patient symptom profiles based on Jaccard distance. Points are coloured by most probable latent class from the five-class LCA model and axes indicate scale in unitless Jaccard distance. The embedding demonstrates substantial overlap between classes and smooth variation in symptom similarity, with no clear separation between categories.

The best-fitting latent trait model had four dimensions. After varimax rotation, the symptoms with the greatest change in probability associated with movement along each of the four dimensions can be summarised as: (1) weakness, gait disorder, and somatosensory symptoms; (2) speech disturbance; (3) depersonalization/derealisation and dystonia; (4) tinnitus, elimination problems, and visual symptoms. Plots of the ten symptoms with the greatest change in probability associated with each dimension, as well as the ten symptoms with the greatest factor loadings for each dimension, are provided in Supplementary Figure 1. Note that because latent dimensions are rotationally non-identifiable, these patterns should be interpreted as descriptive summaries of symptom co-variation rather than unique biological axes.

## Discussion

Using symptom data from a large cohort of people with functional neurological disorder (FND), we examined whether patterns of symptom reporting were more consistent with a single disorder with heterogeneous expression or with multiple symptom-defined disease entities. Across all model comparisons, latent trait models—representing a single underlying entity varying along continuous dimensions—provided a reliably better account of the data than latent class models representing multiple discrete entities. Further, secondary analyses of the best-fitting latent class model showed a low effective number of classes, substantial posterior uncertainty at class boundaries, and extensive overlap in low-dimensional symptom embeddings. Together, these findings indicate that the apparent classes identified by latent class analysis are better interpreted as discretisations of underlying continuous variation rather than as separable subgroups. At the level of symptom phenotype, the data are therefore most compatible with FND representing a single, heterogeneous disorder.

It is important to be explicit about the level of inference supported by these results. Our conclusions concern the structure of FND as inferred from symptom distributions, rather than the number or nature of causal mechanisms. Nonetheless, symptom structure provides an empirical constraint on models of disease organisation. If FND comprised multiple symptom-defined disease entities with distinct mechanisms, these differences would be expected to affect the distribution of symptoms at the population level. The absence of evidence for separable symptom clusters in a dataset of this size (N=697) therefore provides evidence against the hypothesis that FND comprises multiple separable clinical entities. While it remains logically possible that distinct pathophysiological processes could give rise to indistinguishable symptom profiles, such entities would, by definition, be clinically and phenomenologically indistinguishable at present.

Our results are most compatible with the first scenario described in the Introduction, in which FND is a single disease entity with dimensional variation in its presentation. They also constrain the second, intermediate scenario we presented, where multiple upstream mechanisms converge to a single common pathway. Our results suggest that, if such upstream heterogeneity exists, the common pathway itself is the dominant determinant of the phenotype, with upstream differences acting as risk or inciting factors for entry into the disease state, rather than as processes which can drive distinct phenotypes. Under this interpretation, FND behaves as a single entity, even if multiple processes can cause it.

Several studies have described identifiable subgroups of people with FND based on psychological features.^7–10^ These findings do not contradict our results. Psychological characteristics are not features of FND itself, and are more likely to represent risk factors, modifiers, or correlates of the condition. The existence of these psychological subtypes may indicate different psychological risk factors for the development of FND, or even different causal processes leading to a single common pathway, but is compatible with a single underlying pathophysiological process leading to the symptoms of the condition.

In addition, a recent study used LCA to look for symptom clusters within a cohort of 155 children with FND.^11^ The authors divided symptoms into four subgroups, and found that, based on which subgroups of symptoms participants experienced, the best fitting LCA model had two categories. As their intent was solely to describe the clinical presentations seen, they did not compare their models with models of dimensional variation, meaning that their findings can neither support nor refute ours.

Our study has limitations. Symptom data were self-reported and binary, which might obscure differences in symptom severity. The analysis was cross-sectional and based solely on symptom presence, and thus cannot address whether distinct entities might be identifiable using biomarkers or longitudinal trajectories. However, these limitations would be expected to bias the analysis toward identifying categorical subgroups rather than continuous variation. Our central conclusion, which is based on the observation that dimensional models provided a superior account of the data, is therefore strengthened, rather than weakened by these limitations. At present, in the absence of reliable biomarkers for FND, symptom patterns remain the most direct observable consequence of underlying disease structure.

In conclusion, we have shown that the patterns of symptoms described by people with FND are most consistent with the view that FND represents a single entity with dimensionally heterogeneous phenotype. If multiple causal processes contribute to disease development, our results suggest that the generation of its phenotype is dominated by a common mechanism. These findings argue against symptom-defined subtypes of FND and suggest that future efforts to stratify patients may be more productive if they focus on dimensional variation, risk and causal factors, longitudinal course, and treatment response rather than categorical symptom clusters.

## Funding

This work was supported by the Canadian Institute for Advanced Research (CIFAR). JBM was supported by a National Health and Medical Research Council (Australia) Investigator Grant (2010141).

## Competing Interests

D.D.G.P is the unpaid project lead at FND Research Connect.

M.J.E. does medical expert reporting in personal injury and clinical negligence cases. M.J.E. has shares in Brain & Mind, which provides neuropsychiatric and neurological rehabilitation in the independent medical sector. M.J.E. has received financial support for lectures from the International Parkinson’s and Movement Disorders Society and the FND Society (FNDS). M.J.E. receives royalties from Oxford University Press for his book The Oxford Specialist Handbook of Parkinson’s Disease and Other Movement Disorder. M.J.E has received honoraria for medical advice to Teva Pharmaceuticals and educational events. M.J.E. receives grant funding from the National Institute for Health and Care Research (NIHR). M.J.E. is an associate editor of the European Journal of Neurology. M.J.E is a board member of the FNDS. M.J.E. is on the medical advisory boards of the charities FND Hope UK and the British Association of Performing Arts Medicine.

## Notes

### Author Declarations

Ethical approval was granted by the University of Queensland's Human Research Ethics Committee with reference 2023/HE001397.

## References

1. Finkelstein SA, Diamond C, Carson A, Stone J. Incidence and prevalence of functional neurological disorder: a systematic review. J Neurol Neurosurg Psychiatry. Published online December 11, 2024. doi:10.1136/jnnp-2024-334767

2. Hallett M, Aybek S, Dworetzky BA, McWhirter L, Staab JP, Stone J. Functional neurological disorder: new subtypes and shared mechanisms. Lancet Neurol. 2022;21(6):537–550. doi:10.1016/S1474-4422(21)00422-1

3. Mammì A, Harper K, Gray C, Reuber M, Brown RJ. Psychological heterogeneity in functional neurological disorders: A systematic review of studies exploring psychopathological sub-types. J Psychosom Res. 2026;200:112473. doi:10.1016/j.jpsychores.2025.112473

4. Coukos R, Krainc D. Key genes and convergent pathogenic mechanisms in Parkinson disease. Nat Rev Neurosci. 2024;25(6):393–413. doi:10.1038/s41583-024-00812-2

5. Morin S, Legault R, Laliberté F, et al. StepMix: A Python Package for Pseudo-Likelihood Estimation of Generalized Mixture Models with External Variables. J Stat Softw. 2025;113:1–39. doi:10.18637/jss.v113.i08

6. Abril-Pla O, Andreani V, Carroll C, et al. PyMC: a modern, and comprehensive probabilistic programming framework in Python. PeerJ Comput Sci. 2023;9:e1516. doi:10.7717/peerj-cs.1516

7. Cragar DE, Berry DTR, Schmitt FA, Fakhoury TA. Cluster analysis of normal personality traits in patients with psychogenic nonepileptic seizures. Epilepsy Behav EB. 2005;6(4):593–600. doi:10.1016/j.yebeh.2005.03.007

8. Pun P, Frater J, Broughton M, Dob R, Lehn A. Psychological Profiles and Clinical Clusters of Patients Diagnosed With Functional Neurological Disorder. Front Neurol. 2020;11:580267. doi:10.3389/fneur.2020.580267

9. Gilmour GS, Langer LK, Lang AE, MacGillivray L, Lidstone SC. Neuropsychiatric phenotypes in functional movement disorder. CNS Spectr. 2023;28(6):747–755. doi:10.1017/S1092852923002353

10. Mammì A, Harper K, Gray C, Reuber M, Brown RJ. Psychological heterogeneity in functional neurological disorders: A systematic review of studies exploring psychopathological sub-types. J Psychosom Res. 2026;200:112473. doi:10.1016/j.jpsychores.2025.112473

11. Nishi A, Hayakawa I, Okubo Y, Waugh JL, Nariai H. Are symptoms clustered into latent classes in pediatric functional neurological disorder? Epilepsy Behav. 2026;176. doi:10.1016/j.yebeh.2025.110876

