## Supplementary Figure for "The Case Against the ‘S’: Is Functional Neurological Disorder(s) One Condition or Many?"

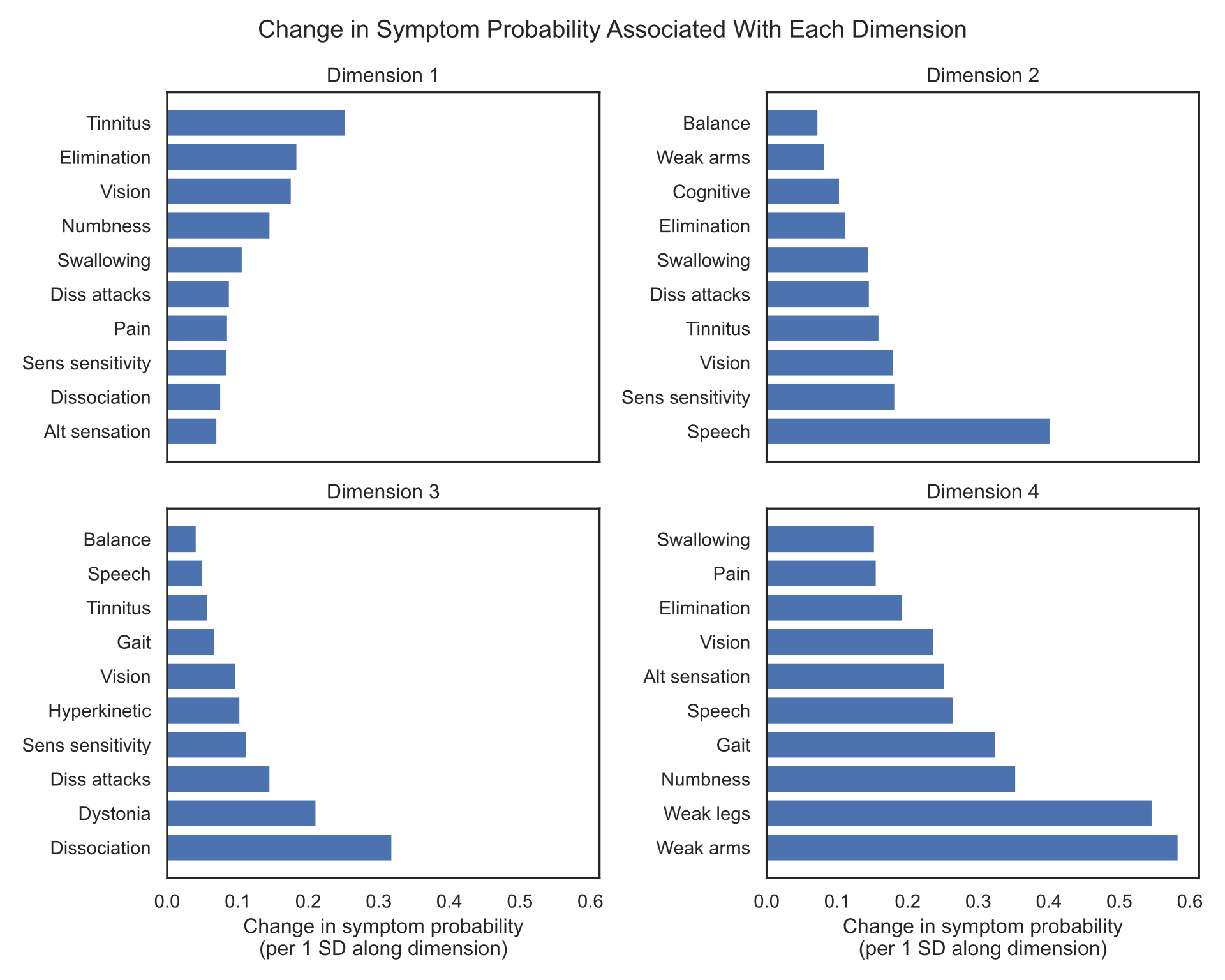

Supplementary Figure 1: Absolute change in symptom probability associated with movement of one standard deviation along each of the dimensions of the four-class latent class model after varimax. Only the ten symptoms with the greatest change in probability for each dimension are presented.

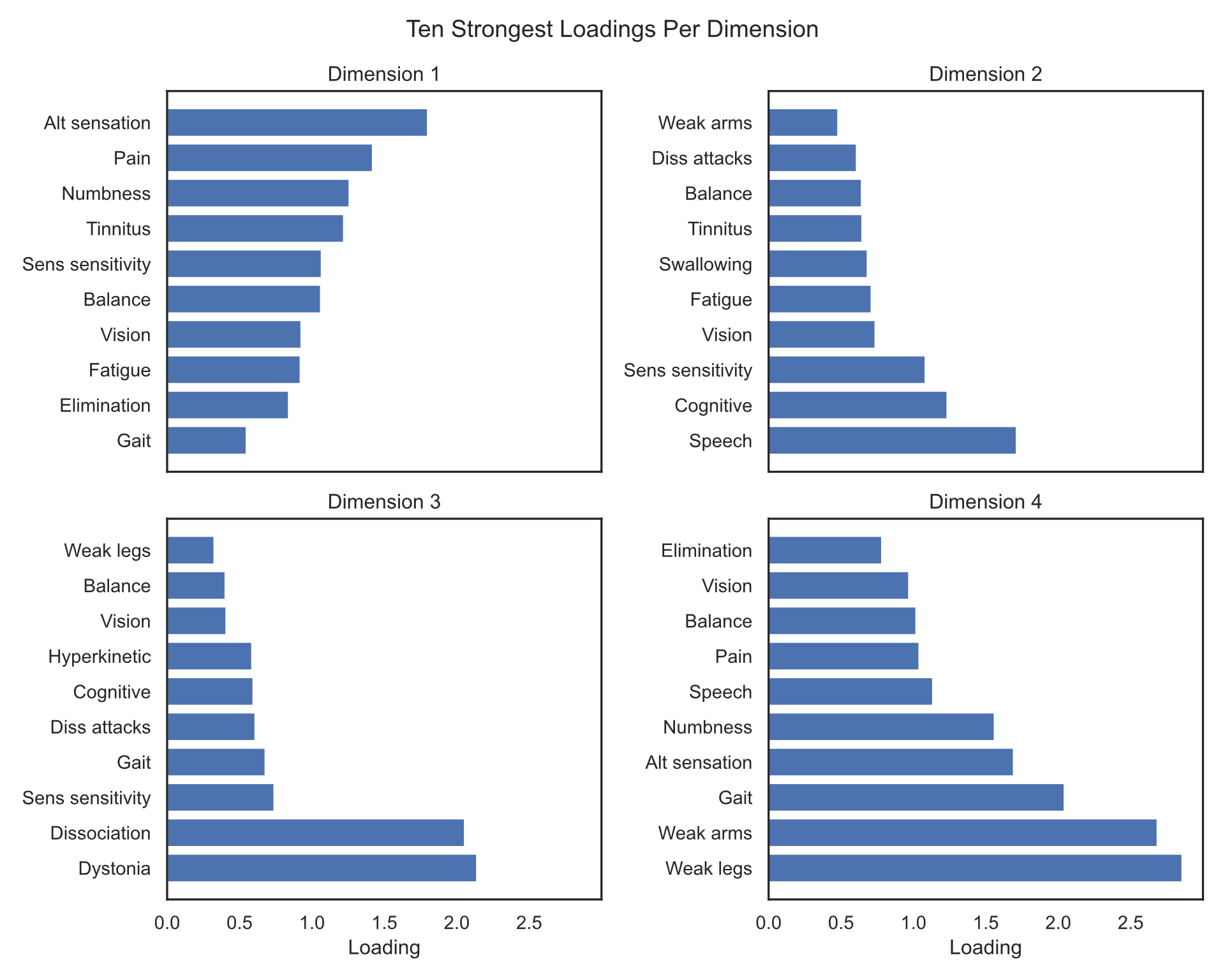

Supplementary Figure 2: Absolute factor loadings of the symptoms with the ten greatest factor loadings for each dimension. Factor loadings indicate how sensitive each symptom is to variation in the underlying latent dimension.
